# Vitamin D status and seroconversion for COVID-19 in UK healthcare workers who isolated for COVID-19 like symptoms during the 2020 pandemic

**DOI:** 10.1101/2020.10.05.20206706

**Authors:** Aduragbemi A Faniyi, Sebastian T Lugg, Sian E Faustini, Craig Webster, Joanne E Duffy, Martin Hewison, Adrian Shields, Peter Nightingale, Alex G Richter, David R Thickett

**Author notes:** Correspondence to Prof David R Thickett, Institute of Inflammation and Ageing, University of Birmingham, Birmingham B15 2TH, UK. Joint first authors (Aduragbemi A Faniyi, Sebastian T Lugg). Joint last authors (Alex G Richter, David R Thickett).

## Abstract

**Background:** It is clear that in UK healthcare workers, COVID-19 infections and deaths were more likely to be in staff who were of BAME origin. This has led to much speculation about the role of vitamin D in healthcare worker COVID-19 infections. We aimed to determine the prevalence of vitamin D deficiency in NHS staff who have isolated with symptoms suggestive of COVID-19 and relate this to vitamin D status.

**Methods:** We recruited NHS healthcare workers between 12^th^ to 22^nd^ May 2020 as part of the COVID-19 convalescent immunity study (COCO). We measured anti-SARS-Cov-2 antibodies using a combined IgG, IgA and IgM ELISA (The Binding Site). Vitamin D status was determined by measurement of serum 25(OH)D_3_ using the AB SCIEX Triple Quad 4500 mass spectrometry system.

**Findings:** Of the 392 NHS healthcare workers, 214 (55%) had seroconverted for COVID-19. A total of 61 (15.6%) members of staff were vitamin D deficient (<30 nmol/l) with significantly more staff from BAME backgrounds or in a junior doctor role being deficient. Vitamin D levels were lower in those who were younger, had a higher BMI (>30 kg/m^2^), and were male. Multivariate analysis revealed that BAME and COVID-19 seroconversion were independent predictors of vitamin D deficiency. Staff who were vitamin D deficient were more likely to self-report symptoms of body aches and pains but importantly not the respiratory symptoms of cough and breathlessness. Vitamin D levels were lower in those COVID-19 positive staff who reported fever, but this did not reach statistical significance. Within the whole cohort there was an increase in seroconversion in staff with vitamin D deficiency compared to those without vitamin D deficiency (n=44/61, 72% *vs* n=170/331, 51%; p=0·003); this was particularly marked in the proportion of BAME males who were vitamin D deficient compared to non-vitamin D deficient BAME males (n=17/18, 94% *vs* n=12/23, 52%; p=0·005). Multivariate analysis revealed that vitamin D deficiency was an independent risk factor for seroconversion (OR 2·6, 95%CI 1·41–4·80; p=0·002).

**Interpretation:** In those healthcare workers who have isolated due to symptoms of COVID-19, those of BAME ethnicity are at the highest risk of vitamin D deficiency. Vitamin D deficiency is a risk factor for COVID-19 seroconversion for NHS healthcare workers especially in BAME male staff.

**Funding:** This study was funded internally by the University of Birmingham and University Hospitals Birmingham NHS Foundation Trust and supported by the National Institute for Health Research (NIHR)/Wellcome Trust Birmingham Clinical Research Facility. AAF and DRT are funded by the Medical Research Council (MR/S002782/1). The Binding Site (Edgbaston, UK) have provided reagents and plates for the SARS-CoV-2 ELISA free of charge.

**Research in context:** *Evidence before this study:* The ongoing COVID-19 pandemic has raised several questions, one of which is whether individuals with vitamin D deficiency were at a greater risk of being infected or having a severe outcome if infected. Among UK healthcare workers, and indeed the general population, individuals of BAME ethnicity are disproportionately affected by COVID-19. It is well established that individuals of BAME ethnicity have a higher prevalence of vitamin D deficiency, but it is unknown if vitamin D deficiency among UK NHS workers was connected to the risk of COVID-19 infection. Our search of the literature revealed no previous studies have established the prevalence of vitamin D deficiency within a UK NHS trust. Unsurprisingly, there is also no evidence to suggest if vitamin D deficiency was connected to the risk of infection among UK healthcare workers.

*Added value of this study:* In this study of healthcare workers who had isolated for COVID-19 symptoms towards the end of UK surge within a large UK NHS trust, 15.6% were vitamin D deficient. Our data also reveal that healthcare workers of BAME ethnicity and those who had seroconverted for COVID-19 were more likely to be vitamin D deficient. Multivariate analysis also show that vitamin D deficiency was the only predictor of COVID-19 seroconversion. Vitamin D deficient healthcare workers that are BAME and male had a 94% seroconversion for COVID-19 compared to non-deficient BAME males suggesting they are more at risk of COVID-19 if vitamin D deficient.

*Implications of all the available evidence:* There is an increased risk of COVID-19 infection in healthcare workers with vitamin D deficiency. Our data adds to the emerging evidence from studies in the UK and across the globe that individuals with severe COVID-19 are more vitamin D deficient than those with mild disease. Finally, ours and the available evidence demonstrate vitamin D supplementation in individuals at risk of vitamin D deficiency or shown to be deficient may help alleviate the impact of COVID-19.

## Introduction

The Severe Acute Respiratory Syndrome Coronavirus 2 (SARS-CoV-2 or COVID-19) pandemic is a global health emergency which has resulted in over 34 million infections, more than 1 million deaths as of the beginning of October 2020,^1^ and is causing a severe global recession.

In vitamin D_3_ deficiency (VDD), immunity becomes dysregulated with phenotypic changes in immune cells particularly lymphocytes and monocytes,^2^ which manifests clinically as an increased susceptibility to infections. In bacterial sepsis vitamin D deficiency is highly prevalent and is a risk factor for the development of the resultant acute respiratory distress syndrome (ARDS).^3,4^ Most patients who die in the intensive care unit (ITU) from COVID-19 have ARDS.^5^ Importantly, the development of a critical illness induces vitamin D deficiency possibly due to dysregulated metabolism.^6^

There has been much speculation about the role of VDD as a determinant of developing symptomatic SARS-CoV-2 infection and whether replacement therapy could be either an effective preventative strategy or even a treatment for those with acute COVID-19 pneumonia.^7^

Data suggest COVID-19 has disproportionately affected those from Black, Asian, and minority ethnic (BAME) groups after accounting for age, sex, social deprivation, and co-morbidity;^8^ VDD is common in this group. Equally the case fatality rate for COVID-19 increases with latitude from the equator,^9^ in common with seasonal influenza and the swine flu pandemic suggesting that sunlight exposure might be important.

During the pandemic it became clear that many healthcare workers were at a higher risk of COVID-19 infection and that in UK healthcare workers deaths were more likely to be in staff who were of BAME origin and particularly those born abroad.^10^ This has led to much speculation about the role of vitamin D in healthcare worker COVID-19 infections.

The prevalence of vitamin D deficiency in healthcare workers in the NHS has not been widely studied. Studies looking at healthcare workers outside the UK suggest that shift workers are more deficient than daytime workers and in addition, looking at medical staff, junior doctors (or residents) were more deficient than senior doctors (or practising physicians).^11^ Given that the COVID-19 pandemic hit at the end of the winter when vitamin D levels are lowest, we sought to address the following aims with this study.

What is the prevalence of vitamin D deficiency in NHS workers who have isolated for symptoms during COVID-19? What were the demographic and occupational determinants of VDD in the NHS healthcare cohort? How did this relate to self-reported symptoms? Was there a relationship between COVID-19 infection and vitamin D status?

## Methods

### Participant recruitment

This prospective observational study recruited healthcare workers between 12^th^ to 22^nd^ May 2020 from the University Hospitals Birmingham NHS Foundation Trust (UHBFT) across four sites as part of the COVID-19 convalescent immunity study (COCO). The study was approved by the London - Camden & Kings Cross Research Ethics Committee (20/HRA/1817). Email adverts were sent to hospital staff to inform them of the study with written informed consent obtained for all participants. The main inclusion criteria were that staff members have had symptoms suggestive of COVID-19. Participants were also asked to provide demographic details such as age, BMI, sex, ethnicity, job role, and co-morbidities as well as clinical details such as details of COVID-19 illness and symptoms. After obtaining consent, blood samples were taken which were immediately transported safely to the laboratory for processing to obtain serum for SARS-CoV-2 antibody and vitamin D assay. All sample processing was done at Biosafety level 2.

### SARS-CoV-2 antibody assay

We measured anti-SARS-CoV-2 antibodies using an in-house IgG, IgA, IgM combined ELISA antibody previously reported.^12,13^ The ELISA is CE-marked with 98.3% (95% CI: 96.4-99.4%) specificity and 98.6% sensitivity (95% CI: 92.6-100%).^14^ Briefly, high-binding plates (Nunc-Maxi-sorp) were coated with 1 μg/ml trimeric SARS-CoV-2 spike glycoprotein,^15,16^ and blocked with Stabil coat solution (Sigma Aldrich). This was followed by addition of pre-diluted serum (1:40 dilution). Antibodies were detected using a combined secondary layer containing horse-radish peroxidase conjugated polyclonal antibodies against IgG, IgA and IgM (The Binding Site, UK). Plates were developed using TMB core (The Binding Site) with orthophosphoric acid (The Binding Site) used as stop solution. Optical densities at 450_nm_ (OD_450nm_) was measured using the Dynex Revelation automated liquid handler. Samples with mean OD_450nm_ plus 2 standard deviations (+2SD) above pre-2019 negative serum control samples were reported as positive for anti-SARS-CoV-2 antibodies. We have used seroconversion as a marker of infection to indicate if an individual has been previously infected by SARS-CoV-2.

### Vitamin D assay

Vitamin D status was determined by measurement of serum 25(OH)D_3_. Serum samples were subjected to a protein crash followed by online extraction and 25(OH)D_3_ was quantified using LC-MS/MS, specifically a Shimadzu UPLC system with an AB SCIEX Triple Quad 4500 mass spectrometer. For this study participants were classified as either vitamin D deficient if serum 25(OH)D_3_ concentration is below 30 nmol/l or as not deficient if it is greater than or equal to 30 nmol/l.^17^ The threshold for vitamin D deficiency varies across studies and regions. However, we have used less than 30 nmol/l as deficient based on the UHBFT clinical laboratory reference guidelines which are in line with UK National Osteoporosis Society guideline for vitamin D.^18^

### Statistics

Results are expressed as median (IQR) for non-normally distributed continuous variables and as a percentage for categorical variables. The Mann Whitney U test and Fisher’s exact test were used for comparison between two groups and contingency tables respectively. Comparison between multiple groups was done by Kruskal-Wallis analysis followed by Dunn’s test. A backward multivariate binary logistic regression analysis was performed to identify independent demographic and occupational factors associated with vitamin D deficiency and seroconversion within this dataset. All analysis was performed using the IBM SPSS V.25 and GraphPad Prism V.8. A P value less than 0.05 was determined as significant.

### Sample size

The COCO staff study was an urgent study to assess convalescent immunity in NHS staff at the University Hospital Birmingham NHS Foundation trust. As the prevalence of seroconversion or staff vitamin D levels was unknown, no formal sample size calculation was possible prior to the study. An amendment to the main ethics for inclusion of vitamin D measurement was approved on the 3rd of May 2020. A total of 460 healthcare workers were recruited to the COCO study after the amendment. Nine participants were excluded from the analysis as their SARS-Cov-2 antibody results were equivocal and a further 59 patients were excluded as they had isolated only due to household members being ill and were asymptomatic. Hence, a total of 392 participants were included in the vitamin D analysis. This included 214 (55%) who had seroconverted and 178 (45%) who had not seroconverted.

### Role of funding source

The study design and views expressed in this paper are those of the authors and it was not influenced by the funding sources. Research support was provided by the National Institute for Health Research (NIHR)/Wellcome Trust Birmingham Clinical Research Facility. Laboratory work was done at the Clinical Immunology Service of the University of Birmingham and the Biochemistry department within the University Hospitals Birmingham NHS Foundation Trust.

## Results

Of the 392 healthcare workers studied, a total of 61 (15·6%) were vitamin D deficient. The serum 25(OH)D_3_ levels of the whole cohort were 55·5 (IQR 39·3–69·1) nmol/l, with levels of 22·0 (15·7– 26·0) nmol/l in the vitamin D deficient group and 59·2 (IQR 46·5–73·2) nmol/l in the non-deficient group (p<0·001). The participant demographics and occupation are displayed in Table 1. The median age of the cohort was 41 years (IQR 30–50), 285 (73%) were female, 279 (71%) were white ethnicity and the median BMI was 25·9 (22·9–30·1) kg/m^2^. Of the cohort 240 (61%) had no co-morbidities.

**Table 1:** Participant demographic, occupation and seroconversion status

|  | **Total**<br>**(n=392)<sup>a</sup>** | **Vitamin D**<br>**deficient**<br>**(n = 61)<sup>a,b</sup>** | **Non-Vitamin D**<br>**deficient**<br>**(n = 331)<sup>a,b</sup>** | **p value** |
| --- | --- | --- | --- | --- |
| Age years, median (IQR) | 41 (30–50) | 35 (28–47·5) | 42 (31–50) | 0·073 |
| Gender no. (%) |  |  |  |  |
| Female | 285 (73%) | 40 (66%) | 245 (74%) | 0·112 |
| Male | 100 (26%) | 21 (34%) | 79 (24%) |  |
| Not stated | 7 (2%) | .. | 7 (2%) |  |
| BMI kg/m<sup>2</sup>, median (IQR) | 25·9 (22·9–30·1) | 25·4 (22·9–30·8) | 26·0 (22·9–30·1) | 0·794 |
| Ethnicity no. (%) |  |  |  |  |
| White | 279 (71%) | 18 (30%) | 261 (79%) | <0·0001 |
| BAME | 108 (28%) | 43 (70%) | 65 (20%) |  |
| Not stated | 5 (1%) | .. | 5 (2%) |  |
| Co-morbidities no. (%) |  |  |  |  |
| None | 240 (61%) | 40 (66%) | 200 (60%) | 0·478 |
| One or more | 152 (39%) | 21 (34%) | 131 (40%) |  |
| Job role no. (%) |  |  |  |  |
| Junior doctor | 50 (13%) | 15 (25%) | 35 (11%) | 0·029<sup>c</sup> |
| Consultant | 65 (17%) | 7 (11%) | 58 (18%) |  |
| Junior nurse | 65 (17%) | 12 (20%) | 53 (16%) |  |
| Senior nurse | 66 (17%) | 7 (11%) | 59 (18%) |  |
| Physiotherapist | 28 (7%) | 4 (7%) | 24 (7%) |  |
| Laboratory worker | 26 (7%) | 7 (11%) | 19 (6%) |  |
| Radiology/Theatre staff/Pharmacy | 21 (5%) | 1 (2%) | 20 (6%) |  |
| Secretary/Administrator | 35 (9%) | 2 (3%) | 33 (10%) |  |
| Health Care Assistant/Phlebotomist | 36 (9%) | 6 (10%) | 30 (9%) |  |
| Seroconversion |  |  |  |  |
| Yes | 214 (55%) | 44 (72%) | 170 (51%) | 0·003 |
| No | 178 (45%) | 17 (28%) | 161 (49%) |  |

<sup>a</sup> Vitamin D deficient is Serum 25(OH)D<sub>3</sub> < 30 nmol/l while not deficient is ≥ 30 nmol/l; <sup>b</sup> Where proportions are shown, they were calculated using the n numbers shown in columns as denominator; p values were calculated using Mann Whitney test for data showing median and interquartile range (IQR), and by Fisher's exact test for data showing proportions. p value <0·05 is considered significant. <sup>c</sup> P value of 0·226 when excluding junior doctor group in analysis.

On univariate analysis those with vitamin D deficiency were significantly more likely to be in a BAME ethnic group (p<0·0001) and in a junior doctor job role (p=0·029) (Table 1). There was no difference in age, BMI or co-morbidity status between vitamin D deficient and non-deficient groups. Vitamin D levels were however lower in those who were younger, had a higher BMI (>30 kg/m^2^) and were male (Figure 1A-C). Vitamin D levels of junior doctors were significantly lower than consultants (p<0·01) as well as those in a radiology/theatre and pharmacy setting (p<0·01) or senior nurse (p<0·05), or phlebotomy/healthcare assistant role (p<0·05) (Figure 1D).

**Figure 1:**
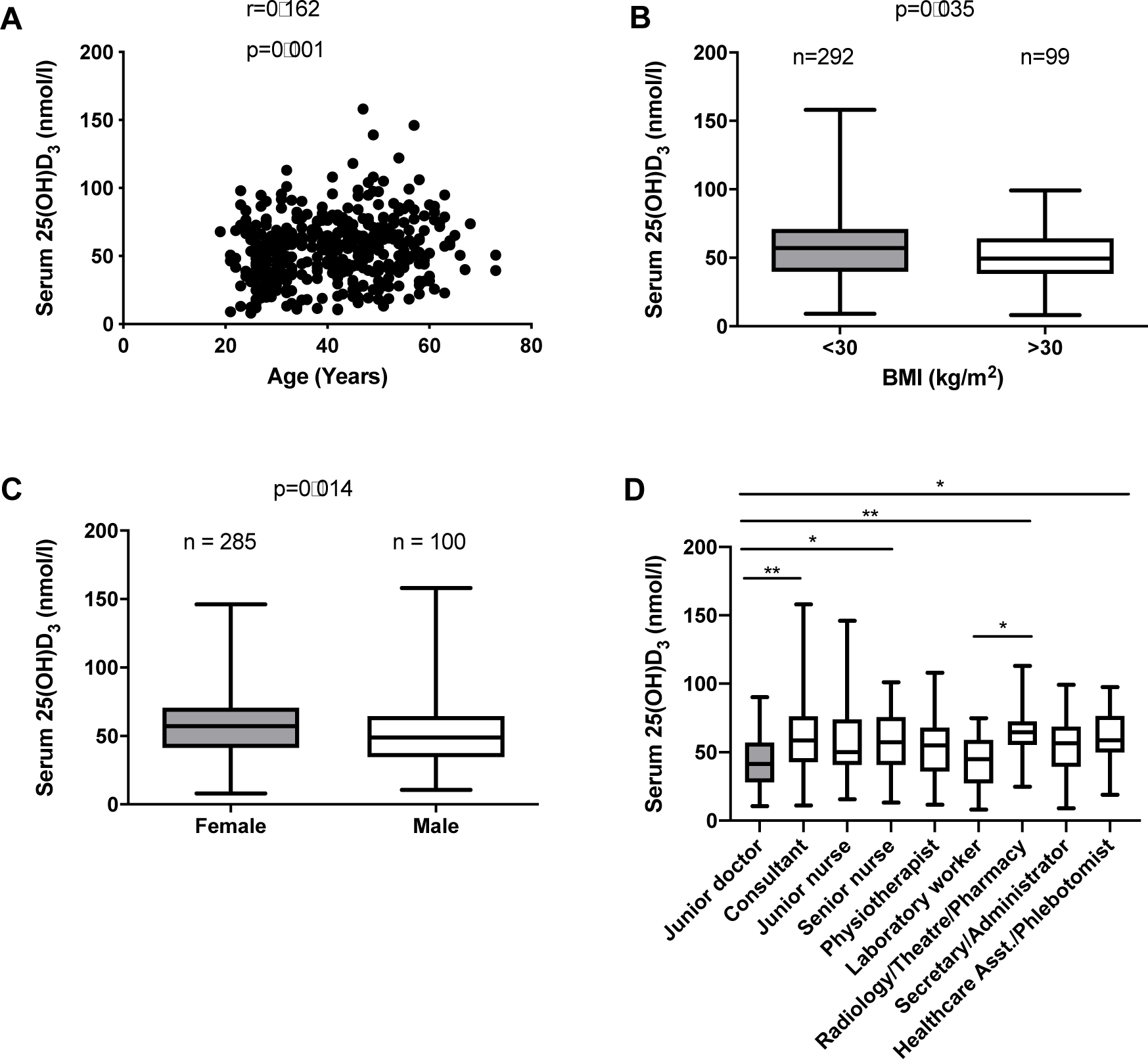
Serum 25(OH)D_3_ concentration in study cohort. **A**. Correlation of vitamin D concentration with age. **B**. Serum 25(OH)D_3_ levels in healthcare workers with BMI less than and greater than 30 kg/m^2^. **C**. Comparison of vitamin D levels in male and female staff while **D** shows the healthcare workers levels of vitamin D according to their job roles. Statistical significance was calculated using spearman correlation (A), Mann Whitney test (B and C), and one-way ANOVA Kruskal-Wallis test with Dunn’s multiple comparison (D). *p<0·05, **p<0·01.

Vitamin D levels were lower in the BAME ethnic group compared to the white ethnic group (p<0·0001) (Figure 2A). In those with Vitamin D deficiency, levels were lower in BAME ethnicity compared to white ethnicity (p=0·001) (Figure 2B), with no significant difference between age and gender between groups. There was, however, no difference in vitamin D levels between BAME and white ethnic groups in those who were non-vitamin D deficient (Figure 2C).

**Figure 2:**
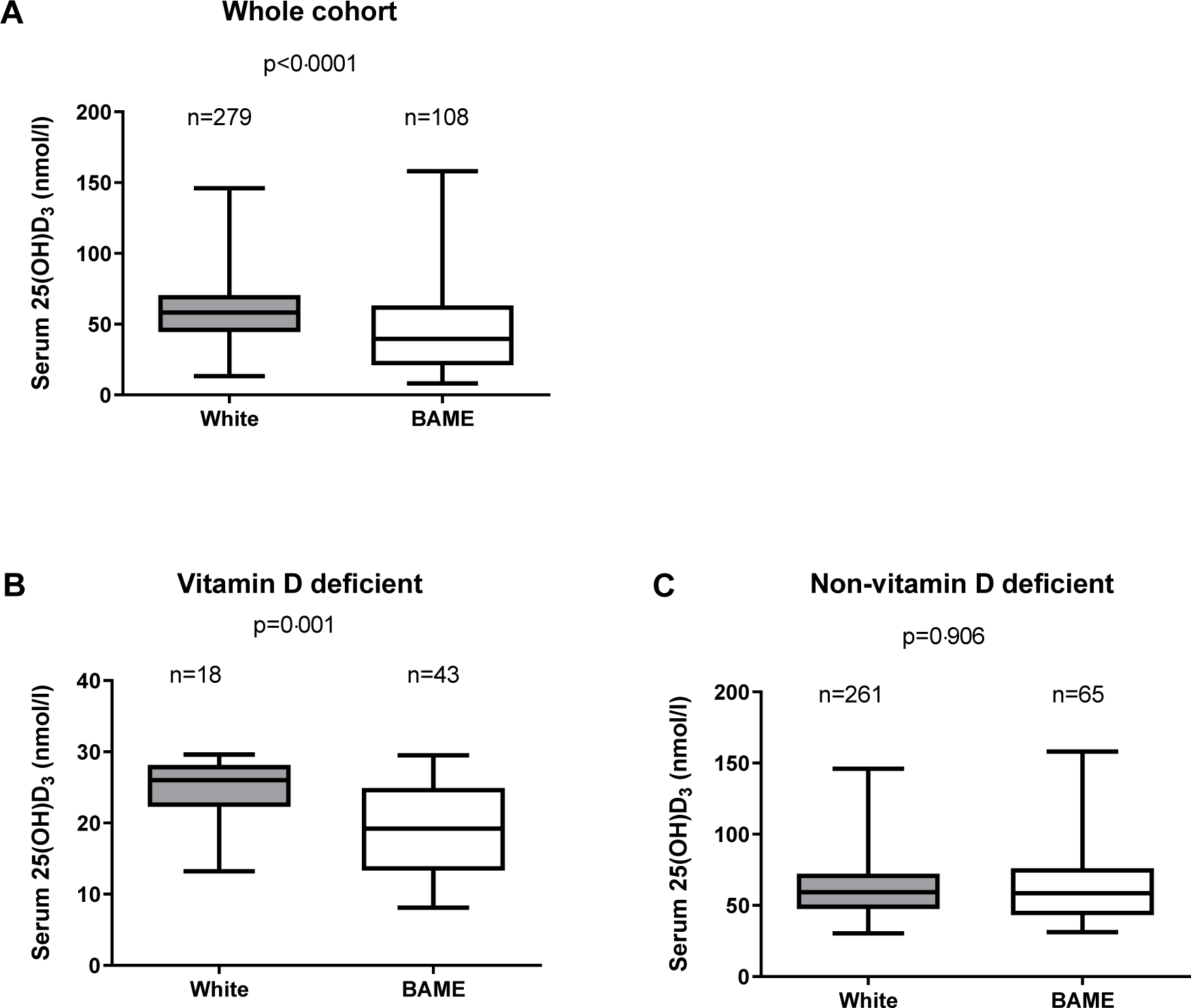
Serum D3 concentration in white and BAME staffs. Vitamin D levels in white and BAME cohort is shown in **A. B** shows healthcare workers that are vitamin D deficient (Serum 25(OH)D_3_ < 30 nmol/l) while **C** represents staffs that are not vitamin D deficient (Serum 25(OH)D_3_ ≥ 30 nmol/l). Mann Whitney test was used to determine statistical significance, p value <0·05 is considered significant.

Using backwards logistic regression to determine factors associated with vitamin D deficiency, the multivariate analysis used included the patient demographic variables of age, gender, BMI, ethnicity, co-morbidities, job role, and seroconversion (Table 2).

**Table 2:**
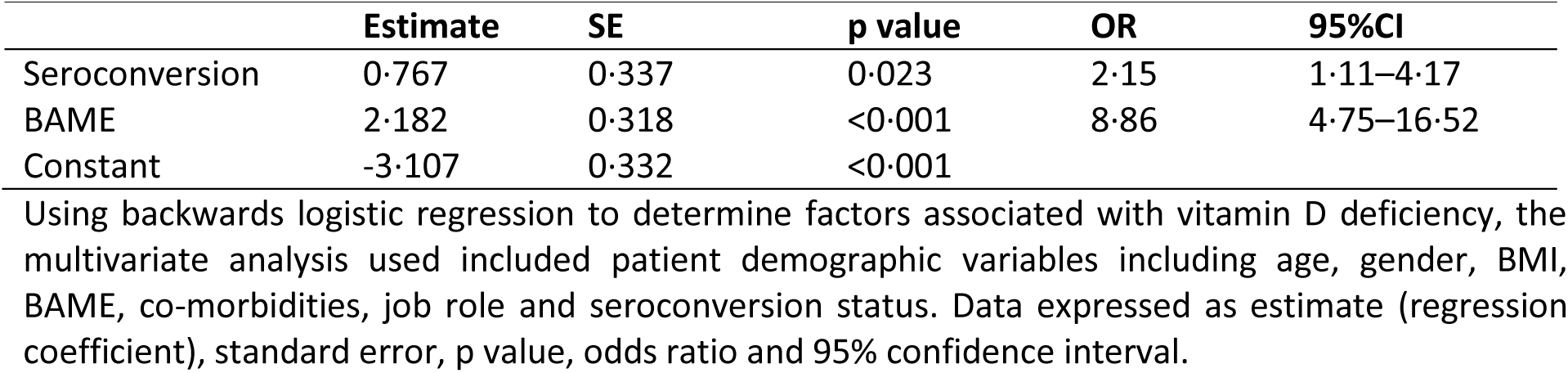
Multivariate analysis of variables related to Vitamin D deficiency

|  | **Estimate** | **SE** | **p value** | **OR** | **95%CI** |
| --- | --- | --- | --- | --- | --- |
| Seroconversion | 0.767 | 0.337 | 0.023 | 2.15 | 1.11–4.17 |
| BAME | 2.182 | 0.318 | <0.001 | 8.86 | 4.75–16.52 |
| Constant | -3.107 | 0.332 | <0.001 |  |  |

Using backwards logistic regression to determine factors associated with vitamin D deficiency, the multivariate analysis used included patient demographic variables including age, gender, BMI, BAME, co-morbidities, job role and seroconversion status. Data expressed as estimate (regression coefficient), standard error, p value, odds ratio and 95% confidence interval.

The significant independent factors in the model were BAME (OR 8·86, 95%CI 4·75–16·52; p<0·001) and COVID-19 seroconversion (OR 2·15, 95%CI 1·11–4·17; p=0·023). The goodness-of-fit test of this model remained non-significant during these steps, with a p value close to one showing a good fit for the final model (Hosmer and Lemeshow p=0.708). The overall predictive power of the model was 77·9% (95%CI 71·1–84·7, SE 3·5%; p<0.001) indicated by the area under the receiver operator characteristic (ROC) curve. A cut off probability set at 0·185 had a sensitivity of 70·5% and specificity of 81.1%. The predicted probabilities of vitamin D deficiency are included in Table 3.

**Table 3.**
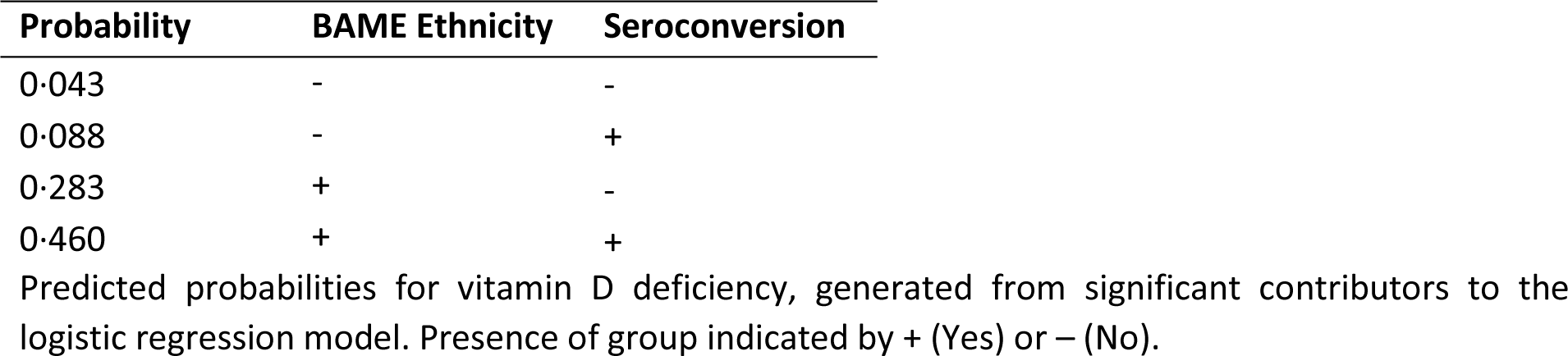
Predicted probabilities for Vitamin D deficiency

| **Probability** | **BAME Ethnicity** | **Seroconversion** |
| --- | --- | --- |
| 0.043 | - | - |
| 0.088 | - | + |
| 0.283 | + | - |
| 0.460 | + | + |

Predicted probabilities for vitamin D deficiency, generated from significant contributors to the logistic regression model. Presence of group indicated by + (Yes) or – (No).

Specific symptoms were not reported by 6 staff, who were all in the non-vitamin D deficient group. Of the patients that self-reported symptoms (n=386), 117 (30%) reported cough, 235 (61%) had fever, 186 (48%) had breathlessness, 169 (44%) had loss of smell or taste, 274 (71%) had body aches and pains, 339 (88%) had fatigue, 115 (30%) had diarrhoea, and 197 (51%) had a sore throat. Staff who had vitamin D deficiency were significantly more likely to experience symptoms of body aches and pains (82% *vs* 69%; p=0·045), but there was however no difference in other symptoms reported between groups (Figure 3A; Supplement Table 1). Vitamin D levels in staffs with body aches and pains were however not different from staffs without the symptom in the whole cohort and those who have seroconverted (Figure 3B). The vitamin D levels however were slightly lower in those who had developed fever within the cohort (p=0·014) and in those who had seroconverted, but this did not reach statistical significance (p=0·055) (Figure 3B).

**Figure 3:**
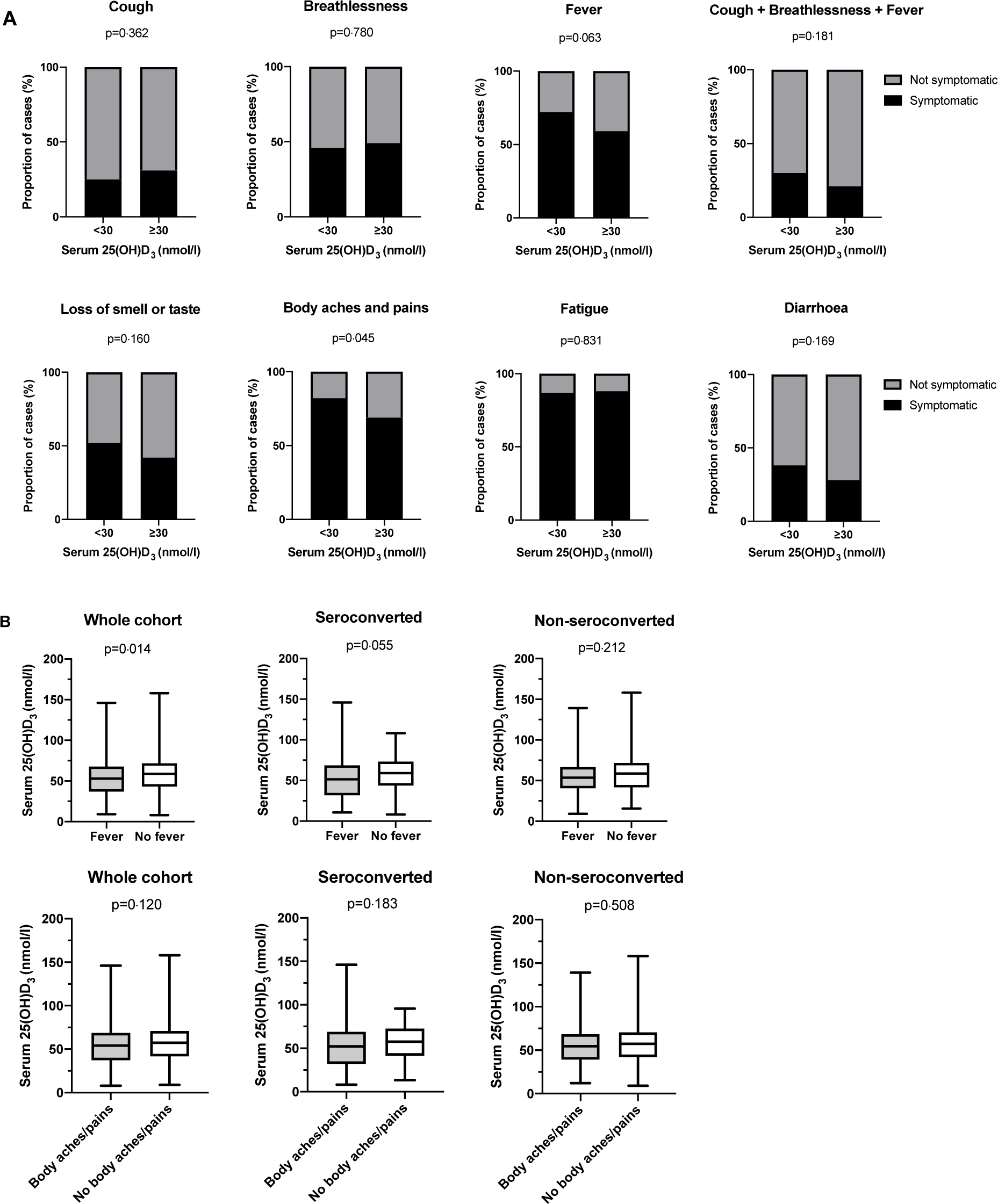
Comparison of symptoms in vitamin D deficient healthcare workers and relationship with seroconversion. **A** Shows the proportion (%) of symptomatic healthcare workers in vitamin D and non-vitamin D deficient groups within the whole cohort. Graphs displayed as percentage of groups, statistical significance was determined using Fisher’s Exact test, p value <0·05 is considered significant. **B** Shows differences of serum vitamin D concentrations in healthcare workers with and without symptoms of fever or body aches/pains within the whole cohort, seroconverted participants and non-seroconverted participants. Graphs displayed as median and IQR; statistical significance was determined using Mann Whitney U test, p value <0·05 is considered significant.

Within the whole cohort there was an increase in seroconversion in healthcare workers with vitamin D deficiency compared to those without vitamin D deficiency (n=44/61, 72% *vs* n=170/331, 51%; p=0·003) (Figure 4A). Overall there was no difference in serum 25(OH)D_3_ levels between seroconverted and seronegative staff (Figure 4B).

**Figure 4:**
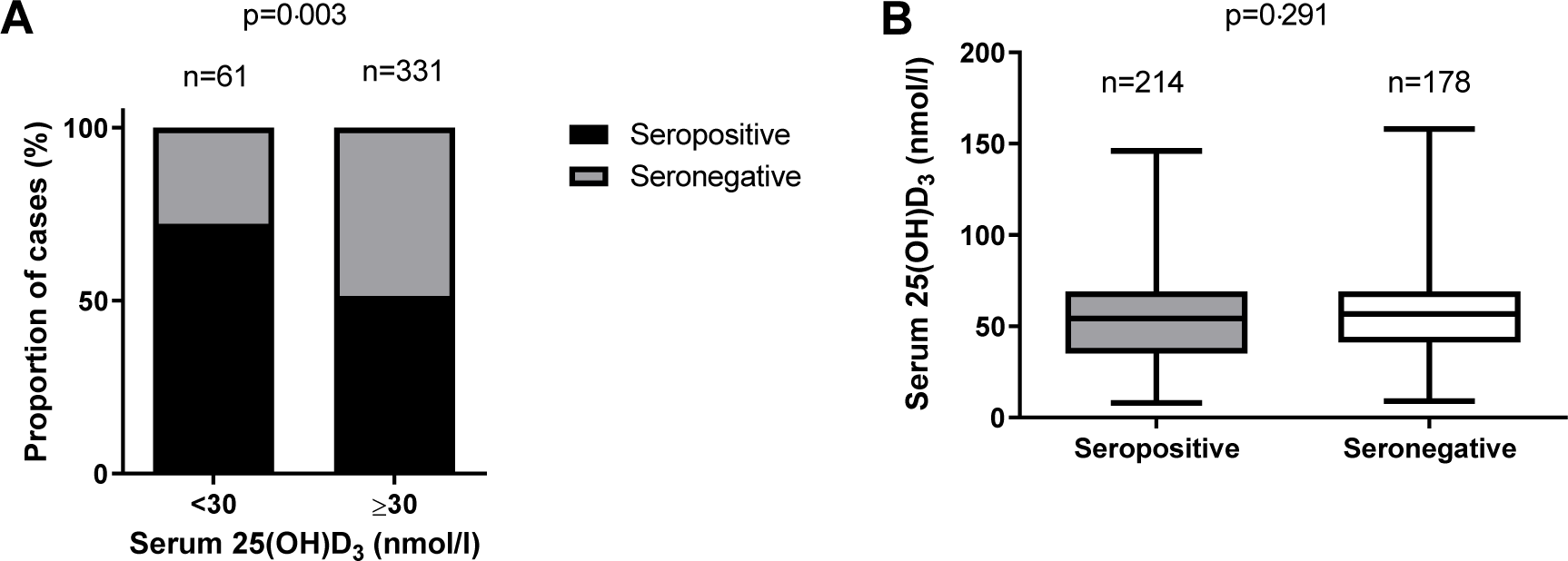
SARS-CoV-2 antibodies seroconversion and vitamin D deficiency. **A** shows proportion of seroconverted healthcare workers in vitamin D deficient and non-deficient groups, while the comparison of serum 25(OH)D_3_ concentration in seropositive and seronegative staffs is shown in **B**. Statistical significance was calculated using Fisher’s exact test (A) and Mann Whitney test (B).

To understand this in more detail the proportion of seroconversion between vitamin D deficient and non-deficient healthcare workers of different sub-groups were assessed (Figure 5A-B). There was no difference in proportion of seroconverted cases between vitamin D deficient and non-deficient healthcare workers of white ethnicity, even after accounting for gender (Figure 5A). Within the BAME ethnic group of the cohort, there was no significant difference in seroconversion between vitamin D status, however, within the BAME male group there was a significant increase in patients who had seroconverted in the vitamin D deficient group compared to the non-deficient group (n=17/18, 94% *vs* n=12/23, 52%; p=0·005) (Figure 5B).

**Figure 5:**
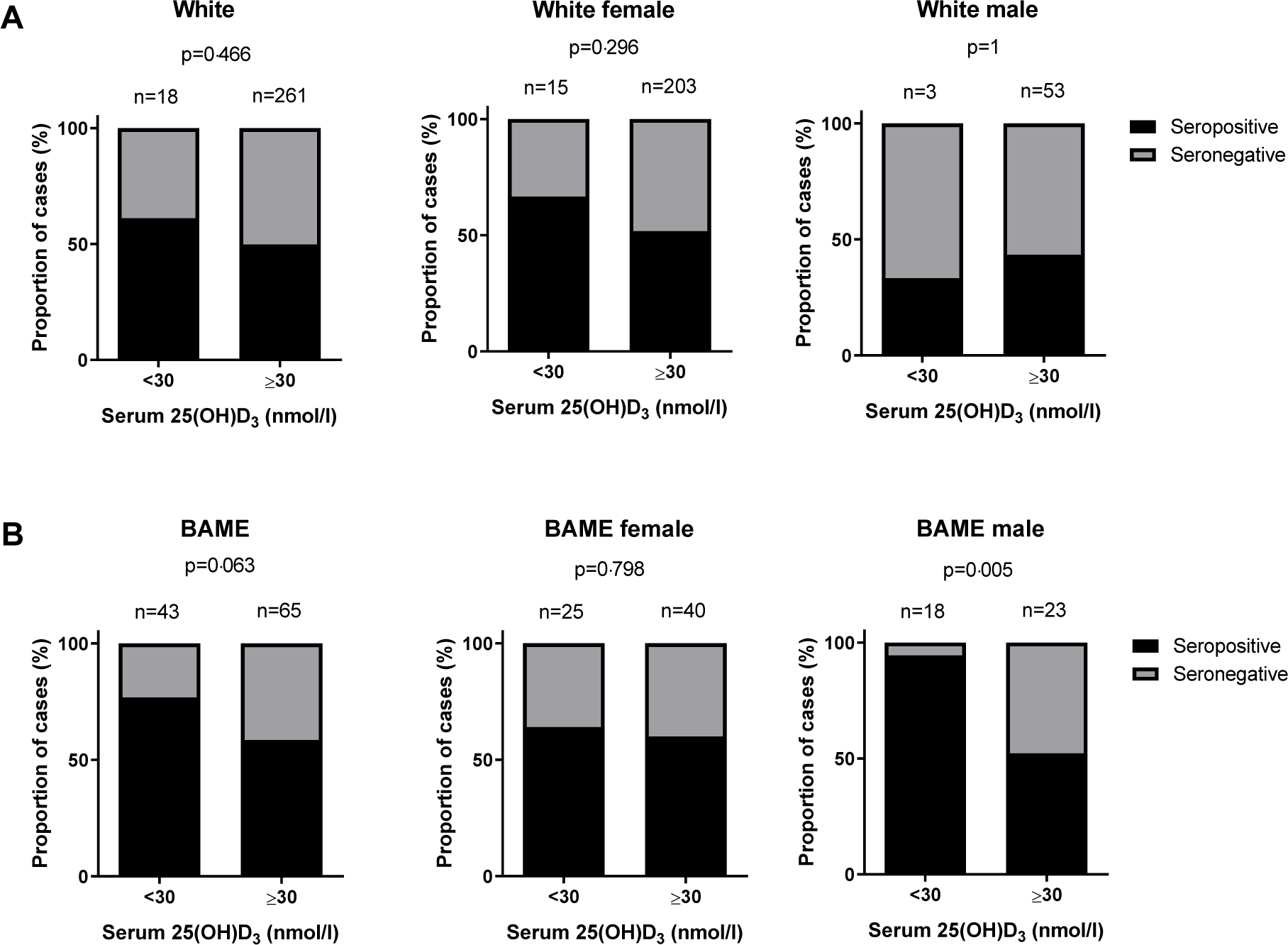
Comparison of seroconversion in vitamin D deficient and not deficient staffs within ethnic subgroups. **A** shows proportion of seroconversion in healthcare workers of white ethnic background including females and males, while **B** also shows breakdown of total, female and male seroconversion in health workers of BAME ethnic background. Statistical significance was determined using Fisher’s exact test, p value <0·05 is considered significant.

Using backwards logistic regression to determine factors associated with seroconversion, the multivariate analysis used included the patient demographic variables of age, gender, BMI, ethnicity, co-morbidities as well as job role and vitamin D deficiency (Table 4). Of those assessed only vitamin D deficiency was a significant independent risk factor for developing seroconversion (OR 2·6, 95%CI 1·41–4·80; p=0·002). The overall predictive power of the model was 55·5% (95%CI 49·8–61·2, SE 2·9%; p=0·06) as indicated by the area under the receiver operator characteristic (ROC) curve.

**Table 4:** Multivariate analysis of variables related to seroconversion

| **Variable** | **Estimate** | **SE** | **p value** | **OR** | **95%CI** |
| --- | --- | --- | --- | --- | --- |
| Vitamin D deficiency | 0.955 | 0.313 | 0.002 | 2.60 | 1.41–4.80 |
| Constant | 0.056 | 0.112 | 0.614 |  |  |

Using backwards logistic regression to determine factors associated with seroconversion, the multivariate analysis used included patient demographic variables including age, gender, BMI, BAME, co-morbidities, job role and vitamin D deficiency (25(OH)D<sub>3</sub> < 30 nmol/l vs ≥30 nmol/l). Data expressed as estimate (regression coefficient), standard error, p value, odds ratio and 95% confidence interval.

## Discussion

In this study we have assessed the vitamin D status of a large cohort of NHS healthcare workers towards the end of the first UK surge in the COVID-19 pandemic; 55% of the cohort had seroconverted for COVID-19. Vitamin D deficiency was detected in 15·6 % of the NHS healthcare workers, with significantly more staff from BAME backgrounds or in a junior doctor role being deficient. Vitamin D levels were lower in those who were younger, had a higher BMI (>30 kg/m^2^), and were male. Multivariate analysis revealed that BAME and COVID-19 seroconversion were independent predictors of vitamin D deficiency. Staff who were vitamin D deficient were more likely to self-report symptoms of body aches and pains but importantly not the respiratory symptoms of cough and breathlessness. Vitamin D levels were lower in those COVID-19 positive staff who reported fever, but this did not reach statistical significance. Vitamin D deficient healthcare workers had an increased seroconversion to COVID-19 compared to those with normal levels. This was particularly marked in BAME males who were vitamin D deficient where 94% had seroconverted compared to 52% in non-deficient BAME males. Using backwards logistic regression to determine factors associated with seroconversion, only vitamin D deficiency was an independent risk factor for seroconversion.

Vitamin D deficiency in this staff cohort was relatively uncommon at 15·6%. This is lower than healthcare worker studies published in the USA and Gulf Areas but this in part may reflect differences in reference ranges and assays used to measure vitamin D.^11^ It is also lower than community reported levels in Birmingham,^19^ where 25% of Birmingham patients attending outpatients were VDD. The finding of lower vitamin D levels in BAME staff is not surprising, but the significantly lower levels seen in junior doctors, who had levels lower than all other profession groups, is a novel finding for the UK. However previous reports outside of the UK have suggested that both healthcare students and junior doctors have lower levels than senior doctors.^11^ The observed low levels in junior doctors was not however associated with an increased seroconversion rate.

We believe that this is the first study to implicate COVID-19 seroconversion as an independent risk factor for VDD. This could either reflect an increased risk of developing COVID-19 disease if you have deficiency, or possibly that COVID-19 may have induced vitamin D deficiency, which could be due to dysregulated metabolism as seen in the critically ill where vitamin D levels can fall rapidly.^6^

There have been reports of people of BAME ethnicity being disproportionately affected by COVID-19, and vitamin D deficiency among people of BAME background is well documented. Our data support previous findings of higher vitamin D deficiency in BAME ethnicity,^20^ with BAME ethnicity also being an independent predictor of vitamin D deficiency in the multivariate analysis. While BAME was not an independent risk factor for seroconversion in this cohort, our analysis of the sub-groups shows that vitamin D deficient BAME male may be the group most at risk from COVID-19 as there was remarkably high seroconversion rate of 94% in this sub-group. Although this is a cohort of mild COVID-19, this finding does support previous report that being BAME and male can increase your chances of a severe outcome from COVID-19 if admitted to ICU.^21^ These data raise the question of whether vitamin D supplementation in vitamin D deficient individuals may help alleviate the impact of SARS-Cov-2 if infected.

Seroconversion was more likely in VDD staff than in non-deficient. Above 30 nmol/l there was no evidence of a dose response effect as the proportion of seroconversion was similar when sub-grouped by quartiles or by insufficient / sufficient levels (data not shown). BAME males were found to have a very high proportion of seroconversion, however on multivariate testing only Vitamin D deficiency came out as an independent determinant of seroconversion.

Our data are consistent with a recently published retrospective observational study from the US of over 190,000 patients with matching serum 25(OH)D in the preceding 12 months, which found that testing positive for COVID-19 was inversely related to vitamin D levels. Their finding remained significant in a multivariate model after adjusting for sex, age, latitude, and ethnicity (adjusted OR 0.984 per ng/ml increment 95%CI 0.983–0.986).^22^ Additional data from a managed care organisation in Israel who had serum 25(OH)D levels checked included 782 (10·1%) who tested positive for COVID-19 and 7,025 (89·9%) who tested negative. Multivariate analysis showed that “low vitamin D” (<30ng/ml or <75nmol/l) had an OR 1.45 (1·08–1·95) for COVID-19 positivity and an OR of 1·95 (0·98–4·85) for hospitalisation due to COVID-19.^23^

As previously described, the role of vitamin D in the response to COVID-19 could be twofold.^7^ The first role is through vitamin D supporting production of antimicrobial peptides in the respiratory epithelium, which would make it less likely be infected with the virus and the subsequent development of COVID-19 symptoms. Evidence from clinical meta-analysis is that VDD supplementation can reduce viral upper respiratory tract infections,^24^ so this effect seems plausible.

Secondly, there may be a role of vitamin D to help reduce the body’s response to established COVID-19 infection. In support of this, a study recently reported that older patients with vitamin D deficiency (Serum 25(OH)D <30 nmol/l) had a higher peak D dimer, a marker of inflammation and blood vessel damage, and required more non-invasive ventilation support than patients that were not deficient.^25^ This systemic effect of SARS-CoV-2 on vitamin D may explain why we observed seroconverted staffs with fever had less vitamin D than those without. Vitamin D, is a precursor to a potent steroid hormone influencing a wide range of cellular pathways in organs that are highly relevant to the effects of critical illness and may exert its beneficial effects on acute inflammation, nosocomial infection, respiratory failure, cardiogenic shock and critical illness myopathy.^26^

VDD has been implicated as a risk factor for the development of the acute respiratory distress syndrome,^3^ which is what kills patients with COVID-19 who need ventilation in ICU. Recent studies in the UK,^27^ Italy,^28^ and South Korea^29^ suggest vitamin D deficiency is higher in patients with severe COVID-19 compared to mild cases. This is in contrast to an earlier report using 10-year biobank vitamin D data to suggest vitamin D deficiency is not associated with COVID-19.^30^ However, further evidence from a recent open labelled clinical trial of calcifediol in hospitalised patients with COVID-19 has shown promise as a therapy for severe illness (CORDOBA study) providing a proof of concept that vitamin D therapy may be useful.^31^

This study has limitations. Firstly, staff were recruited from a single NHS trust based in Birmingham which is the second largest NHS trust in the UK with 4 hospital sites. This study recruited only healthcare workers from secondary care hospital settings. The data presented here would benefit from a large validation cohort of staff from across the full diversity of NHS. There is also a need to try and validate these findings in primary care, care homes and hospice settings.

Secondly, the staff cohort who volunteered had mild COVID-19 disease as only 3 out of the whole COCO study cohort were admitted to hospital due to severe disease. Clearly therefore the findings of this study relate only to mild disease and do not reflect the potential effects of VDD in severe COVID-19 disease. Indeed our own data (unpublished) suggest that patients admitted to hospital with COVID-19 have much lower levels of vitamin D than this staff cohort.

Thirdly, due to relatively low numbers of BAME staff members in this cohort we were unable to analyse the differences between staff from different ethnicities. This may be important as it has been demonstrated for example that COVID-19 patients from Bangladeshi origin have worse outcomes from COVID-19 disease.^8^ Whether these differences reflect relevant changes in vitamin D metabolism, genetics, etc remain unknown.

In conclusion, we have shown that in those healthcare workers who have isolated due to symptoms of COVID-19, those of BAME ethnicity are at the highest risk of vitamin D deficiency. Vitamin D deficiency was a risk factor for development of COVID-19 seroconversion, with the biggest differences in seroconversion seen in the BAME male group. Therefore, as vitamin D deficiency is a potential modifiable risk factor for COVID-19 and vitamin D supplementation is cheap, readily available with very little risk of side effects, this study raises the question as to whether it may reduce the risk of COVID-19 disease. We suggest further vitamin D treatment trials should target such at risk groups within healthcare workers.

## Data Availability

Data are available upon reasonable request.
Proposals should be directed to the corresponding author

## Contributors

AGR and DRT conceptualised the study. AAF, SEF, CW, JED, AS, AGR, and DRT contributed to data acquisition. AAF, STL, PN, and DRT analysed the data. All authors contributed to data interpretation. AAF, STL and, DRT drafted the manuscript. All authors contributed to the review and approval of the final copy of the manuscript.

## Declaration of interests

MH reports personal fees from Thornton Ross, outside the submitted work. All other authors declare no competing interests.

## Acknowledgements

We thank the staff of University Hospitals Birmingham NHS Foundation Trust who kindly volunteered for this study. We would also like to thank the research staff of the Birmingham Wellcome NIHR Clinical Research Facility who undertook the staff facing assessments. We would like to thank colleagues at the Clinical Immunology Service for overseeing recruitment and sample processing. We also thank our colleagues Prof Adrian Martineau, Prof Elizabeth Sapey, Dr Dhruv Parekh, and Prof Jon Rhodes who have given helpful feedback on the study results and manuscript.

## Notes

### Author Declarations

London - Camden & Kings Cross Research Ethics Committee

