## Supplement table 1 for "Vitamin D status and seroconversion for COVID-19 in UK healthcare workers who isolated for COVID-19 like symptoms during the 2020 pandemic"

**Supplement Tables**

**Supplement Table 1** Clinical symptoms of whole cohort

| | **Total** (n=386) | **Vitamin D deficient** (n = 61)[a,b] | **Non-Vitamin D deficient** (n = 325)[a,b] | **P value** |
|---|---|---|---|---|
| Cough no. (%) | | | | |
| Yes | 117 (30%) | 15 (25%) | 102 (31%) | 0·362 |
| No | 269 (70%) | 46 (75%) | 223 (69%) | |
| Fever no. (%) | | | | |
| Yes | 235 (61%) | 44 (72%) | 191 (59%) | 0·063 |
| No | 151 (39%) | 17 (28%) | 134 (41%) | |
| Breathlessness no. (%) | | | | |
| Yes | 186 (48%) | 28 (46%) | 158 (49%) | 0·780 |
| No | 200 (52%) | 33 (54%) | 167 (51%) | |
| Cough + fever + breathlessness no. (%) | | | | |
| Yes | 87 (23%) | 18 (30%) | 69 (21%) | 0·181 |
| No | 299 (77%) | 43 (70%) | 256 (79%) | |
| Loss of smell or taste no. (%) | | | | |
| Yes | 169 (44%) | 32 (52%) | 137 (42%) | 0·160 |
| No | 217 (56%) | 29 (48%) | 188 (58%) | |
| Body aches and pains no. (%) | | | | |
| Yes | 274 (71%) | 50 (82%) | 224 (69%) | 0·045 |
| No | 112 (29%) | 11 (18%) | 101 (31%) | |
| Fatigue no. (%) | | | | |
| Yes | 339 (88%) | 53 (87%) | 286 (88%) | 0·831 |
| No | 47 (12%) | 8 (13%) | 39 (12%) | |
| Diarrhoea no. (%) | | | | |
| Yes | 115 (30%) | 23 (38%) | 92 (28%) | 0·169 |
| No | 271 (70%) | 38 (62%) | 233 (72%) | |
| Sore throat (%) | | | | |
| Yes | 197 (51%) | 37 (61%) | 160 (49%) | 0·124 |
| No | 189 (49%) | 24 (39%) | 165 (51%) | |

[a] Vitamin D deficient is Serum 25(OH)$D_3$ < 30 nmol/l while not deficient is ≥ 30 nmol/l; [b] Where proportions are shown, they were calculated using the n numbers shown in columns as denominator; p values were calculated using Fisher's exact test for data showing proportions, p value <0·05 is considered significant.
